# Unveiling Hidden Threats: Introduction of a Routine Workflow for Label-Free and Non-destructive Detection of Microplastics in Human FFPE Tissue Sections

**DOI:** 10.1101/2025.01.09.24319030

**Authors:** Elisabeth S. Gruber, Verena Karl, Kristina Duswald, Mukund S. Bhamidipalli, Michaela Schlederer, Tanja Limberger, Béla Teleky, Lukas Kenner, Markus Brandstetter

**Affiliations:** Department of General Surgery, Medical University Vienna, Vienna, Austria; Research Center for Non-Destructive Testing (RECENDT), Linz, Austria; Center for Biomarker Research in Medicine (CBmed), Graz, Austria; Division of Experimental and Laboratory Animal Pathology, Department of Pathology Medical, University of Vienna, Vienna, Austria; Center for Biomedical Research and Translational Surgery, Medical University of Vienna, Vienna, Austria; Christian Doppler Laboratory for Applied Metabolomics, Medical University of Vienna, Vienna, Austria; Unit of Laboratory Animal Pathology, University of Veterinary Medicine Vienna, Vienna, Austria; Comprehensive Cancer Center, Medical Unviersity Vienna, Vienna, Austria; Department of Molecular Biology, Umeå University, Umeå, Sweden

**Author notes:** Co-correspondence. Equally contributing senior authors (Lukas Kenner, Markus Brandstetter).

**Keywords:** Microplastic, FFPE, colon, inflammation, optical photothermal infrared spectroscopy

## Abstract

Microplastic (MP) pollution has emerged as a significant environmental and health concern. This study aimed at developing a novel method for detecting MP particles in deparaffinized formalin-fixed paraffin-embedded (FFPE) routine tissue sections of human colon samples. By utilizing mid-infrared photothermal (MIP) microscopy, also known as optical photothermal infrared (OPTIR) spectroscopy, we here aimed to establish a work-flow representing a non-destructive infrared spectroscopic approach for distinct identification and localization of polymers in FFPE colon tissue sections by respecting standardized protocols well integrated into clinical routine. After OPTIR analysis, samples were processed to H&E staining for histopathological analysis at defined regions of interest. By using OPTIR spectroscopy, we succeeded in localizing polyethylene (PE), polystyrene (PS), and polyethylene terephthalate (PET) MPs in distinct regions of interest (21 PE particles, 1 PS particle, 1 PET fiber) and clearly identifying the embedded polymers in the respective colon tissue sections. Subsequent H&E analysis confirmed characteristic histopathological features of inflammation spatially associated with the respective MP polymers. We present here for the first time a diagnostic workflow that has the potential to enhance our understanding of MP accumulation in routine organ tissue slides and allows for exploration of its implications for human health. The observed morphological H&E features in proximity to the identified MPs could possibly indicate a link between MP exposure and colon inflammation.

## INTRODUCTION

Microplastic (MP), defined as plastic particles less than five millimeter in size, have been found in various environmental matrices, including marine ecosystems, freshwater bodies, soil and even the air ^1–4^. Their ubiquitous presence has raised concerns about their potential impact on human health ^5–7^. Recent studies have shown that different sorts of MPs can enter the human body through ingestion and inhalation, potentially accumulating in different organs^8–29^.

Polyethylene (PE), polystyrene (PS), and polyethylene terephthalate (PET) are commonly used plastics for food and beverage packaging ^30^ and pose several potential threats to human health, particularly due to their chemical composition and the way they are used or disposed of. While the risks vary depending on exposure levels and individual susceptibility ^31^, the potential health impacts of these plastics are significant, especially in contexts of prolonged or high-level exposure. They can release harmful chemicals, especially when heated or degraded. PS can leach styrene, a possible carcinogen that may cause nervous system effects, irritation, and other health issues ^32,33^. PE, though generally considered safer, can release additives or byproducts during manufacturing or when exposed to heat ^34^. As PE and PS degrade, they break down into MP particles, which can be ingested through food, water, and air, leading to potential health risks such as inflammation, oxidative stress, and disruption of cellular processes ^35–38^. Further (indirect) health impacts can occur through accumulation of PE and PS in the environment, particularly in oceans, contaminating food sources, such as particularly seafood, introducing toxins into the food chain. Generally regarded as safe for food contact^30^, PET raises concerns when degraded into MP particles entering the human body ^22,24^. Studies suggest that PET can carry harmful chemicals like antimony ^39,40^. While there is no conclusive evidence of serious health effects from PET exposure in typical use, the potential for chemical leaching and long-term health risks remains a topic of ongoing research.

Despite growing concerns about the potential health risks of such MP particles, studying their direct effects on the human body remains a challenge. In particular, it is difficult to detect and localize MP particles in human tissue taken as part of routine medical examinations, such as it is the case for formalin-fixed paraffin-embedded tissue (FFPE) used for histopathological analysis and reporting. The tiny size of the particles, their chemical composition and the limited sensitivity of current analytical methods make detection – without losing information about the exact position – difficult, further complicating research into their long-term health effects.

Infrared spectroscopy is a widely used label-free and non-destructive technique to obtain qualitative and quantitative information of organic and inorganic samples. The mid-infrared (MIR) part of the infrared spectrum is here of particular interest as it provides a (bio-)chemical fingerprint and therefore allows characterization and discrimination of biological structures such as lipids, DNA and proteins, e.g. to distinguish diseased from healthy sections ^41,42^. Furthermore, MIR spectroscopy can be used to detect and identify (M)P particles due to their unique chemical compositions. Due to technological limitations of established MIR spectroscopy methods, the current standard procedure for that kind of analysis involves the chemical or enzymatic digestion of the entire tissue sample to remove any biological components. After filtration of the solution, the remaining (M)P particles are collected on a suitable filter for spectroscopic characterization ^8–17,19–29^. However, tissue digestion results in the complete loss of tissue architecture and hence spatial information of accumulated (M)P particles. However, for deeper clinical insight on uptake and accumulation of MPs, it is crucial to identify the precise spatial particle distribution in the respective tissues. To meet this urgent technical need, the recently introduced optical photothermal infrared (OPTIR) spectroscopy technique was employed on deparaffinized human colon samples. OPTIR spectroscopy is characterized by a series of advances compared to conventional MIR spectroscopy methods, among them a significantly improved lateral resolution (∼500 nm) beyond the classical infrared diffraction limit, the ability to measure in reflection geometry on standard glass sample carriers and the absence of spectral artifacts, which are particularly challenging when small structures are investigated^43^.

Our aim here was to test the ability of the OPTIR method for the detection of MP particles, specifically of PS, PE and PET, which are among the most commonly used polymers for food and beverage packaging^30^. The primary objective of this study was to develop and validate a method for detecting MP particles in human colon tissue sections cut from routine FFPE tissue blocks. Specific aims included: 1) Optimizing sample preparation, 2) utilizing the OPTIR spectroscopy technique for the identification of MP particles in histological sections.

## MATERIALS AND METHODS

### Sample Collection and Preparation

Human colon samples were obtained from surgical resections and underwent standard paraffinization work-up to ensure routine histopathological examination and reporting. For this, the samples were fixed in 10% formalin for 24-48 hours. Following fixation, the tissues were dehydrated through a graded series of ethanol solutions, cleared in xylene, and embedded in paraffin blocks.

Ethical approval for the study was given by the local ethics committee (Medical University Vienna, Vienna, Austria, No. 1003/2024). For MP analysis, sections of 5 µm thickness were cut from paraffin blocks using a microtome and mounted on both standard glass (for OPTIR analysis) and low-e (for laser direct infrared (LDIR) imaging analysis) microscope slides. To limit signal reduction and hence spectral characteristics of tissue samples ^44^, deparaffinization was performed according to an established protocol ^45^: In short, slides were first heated to melt the paraffin. After rehydration through graded isopropyl alcohol solutions, the slides were first analyzed with infrared spectroscopy (OPTIR and/or LDIR) and then stained with hematoxylin and eosin (H&E staining). Partial dehydration was done before eosin application, and the slides were mounted with aqueous glycerol and sealed with nail polish.

### Optical Photothermal Infrared (OPTIR) Spectroscopy

For the acquisition of infrared spectra an OPTIR system (mIRage+R, Photothermal Spectroscopy Corp., USA) was employed. The OPTIR technique utilizes a pump-probe approach where the material-dependent spectral absorption of a mid-infrared laser induces a photothermal effect that is read out by a visible laser. The system uses a widely tunable MIR quantum cascade laser (QCL) to locally excite a sample. When the sample absorbs a photon of a specific MIR wavelength, different photothermal effects are triggered (expansion, refractive index change). A second readout laser (532 nm) then probes the photothermal effects, allowing for the detection of the smallest changes ^46^. Selectivity is obtained by the wavelength selective measurement, whereas sensitivity is obtained by the signal strength, which correlates with the amount of substance present. This approach overcomes the infrared diffraction limit and achieves a spatial resolution improved by a factor of ∼30 when compared to conventional FTIR microscopes ^47^. Thus, small structures such as MP particles embedded in complex tissues can be resolved in the submicron range (∼500 nm). For selected samples, additional large-area overview scans at standard infrared resolution limits using laser direct infrared (LDIR) imaging were performed (8700 LDIR Chemical Imaging System, Agilent Technologies, USA). The purpose of these measurements was a faster recognition of larger MP structures (e.g. polymer fibers). Both OPTIR and LDIR were tailored precisely for application to deparaffinized sections.

### Workflow for spatial localization and chemical identification of microplastic particles

To allow for a clear spatial identification of MP particles, an optimized spectroscopic workflow was elaborated by exemplarily focusing on PE particles (**Fig. 1**). The visible image shows the standard optical microscope image of the sample. The chemical images “tissue” and “PE” are derived from the measured infrared absorption at specific wavelengths that are known to be characteristic for tissue and PE, respectively. In this study, the presence of tissue is visualized by generating chemical images derived from the intensity of the amide I band (1660 cm^-1^), as it is a characteristic marker for proteins and organic tissue. In contrast, chemical imaging for the detection of MPs is conducted at a polymer specific wavelength (e.g. represented by the C-H stretching vibration of PE at 2855 cm^-1^). The final step of image processing is illustrated in the chemical image “PE/tissue”, where the ratio of the chemical image intensities of PE and tissue is calculated. The effect of this approach can be interpreted as background normalization. While distinct structures are not recognizable in the single chemical images, the here conducted ratio calculation effectively highlights three PE particles. As demonstrated in subsequent Fig.s, this technique is well suited to distinctly differentiate polymer structures from the surrounding tissue, based on the spectral information obtained at characteristic wavenumbers (= inverse wavelength).

**Fig. 1.**
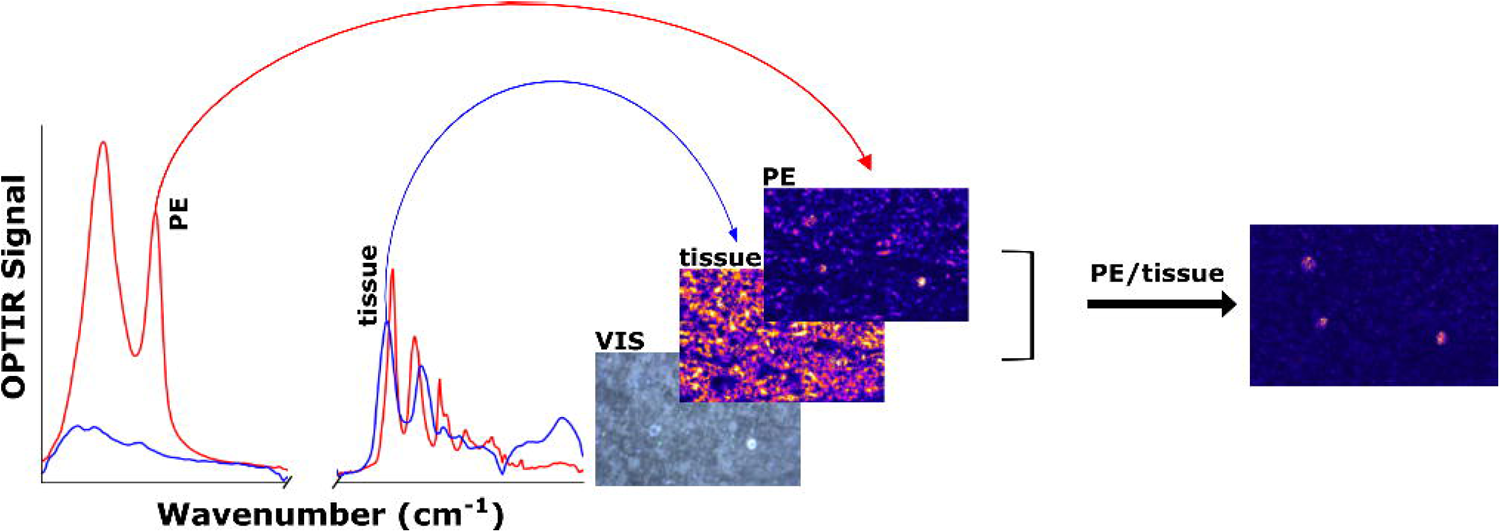
Elaboration of a spectroscopic workflow for optimized MP particle detection. **A.** The visible image (VIS) represents the standard optical microscope image of unstained tissue; **B.** The “tissue”-image shows a chemical image (wavenumber 1660 cm^-1^); **C.** the “PE”-image shows a “chemical” image (wavenumber 2855 cm^-1^) characteristic for the polymer of interest (here: PE). **D.** Final step: “PE/Tissue”-image, calculated as the ratio of “PE”- and “tissue”-image, resulting in a clear identification of PE particles at three different positions within the section/region of interest.

After implementing these technical optimizations, the following “optimized” workflow was defined for the detection of MP particles:

1. Acquisition of visible images of the entire tissue section using the internal optical microscope of the OPTIR system.
2. Selection of distinct regions of interest (region I - mucosa, region II - muscularis, region III - serosa) in the respective colon tissue sections (**Fig. 2**).
3. Chemical Analysis of each region and identification of suspicious particles based on a 2D image recorded at characteristic wavenumbers targeting the polymer type of interest.
4. Validation of the identified PE, PS and/or PET particles based on their characteristic spectral fingerprint (unveiling the chemical nature of the distinct particles by recording full spectra).

**Fig. 2.**
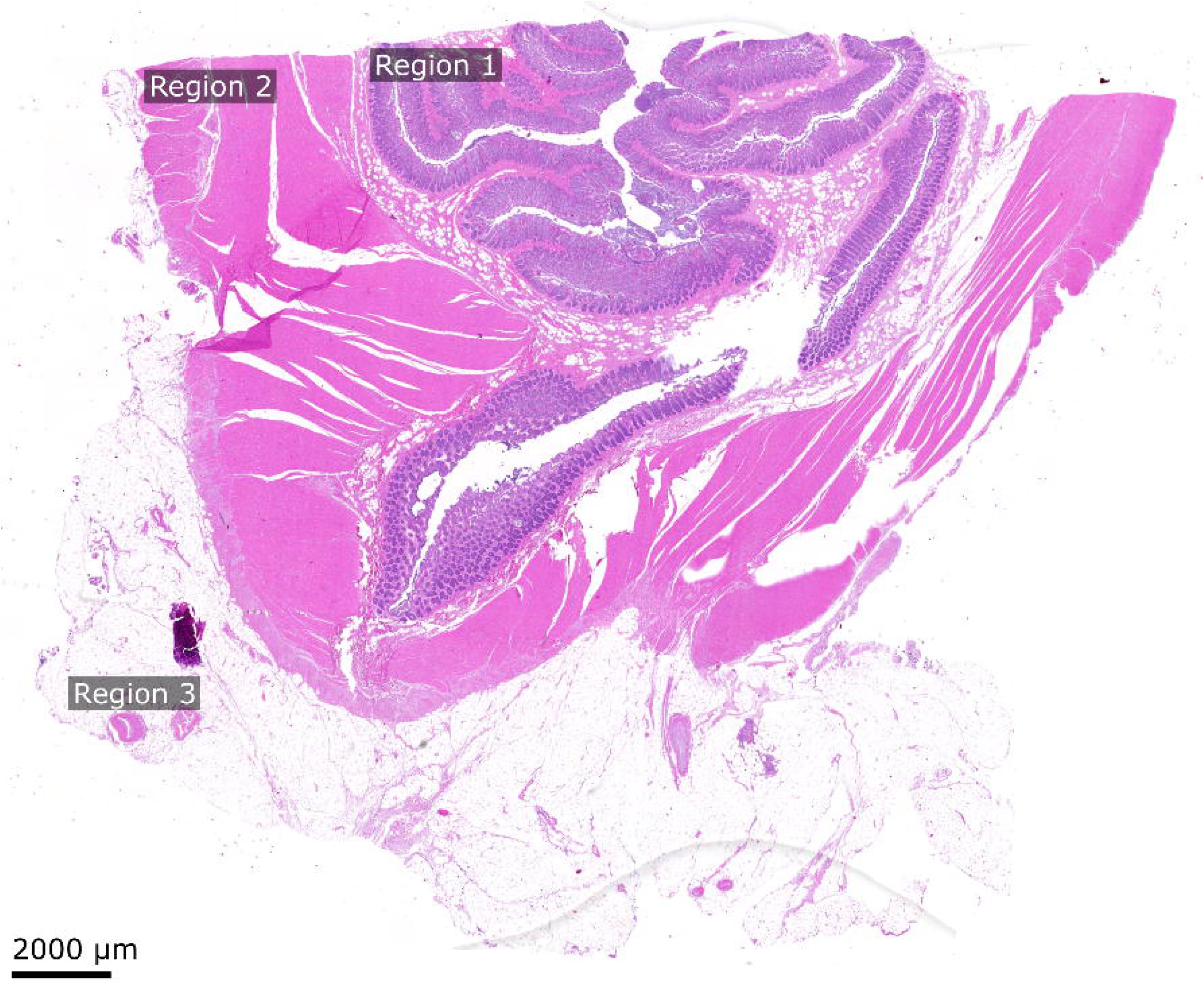
Regions of interest for the exact spatial localization of MP particles. Three regions were defined: region I – mucosa, region II – muscularis and region III – serosa. Since particles are suspected to be absorbed via the mucosa, the primary focus was set to region I and II, which were then analogously inspected for suspicious particles.

Of note, this workflow is generic and can be extended to any other polymer type by simply changing the selected target wavenumbers in step 3.

## RESULTS & DISCUSSION

According to the evident accumulation of MP particles in human organ tissues ^8–29^, the need for further research into potential health threats is of utmost importance. Although several experimental approaches have been introduced ^48^, the clear identification of MP particles remains a technical challenge.

By using OPTIR, we have developed a reliable method for detecting PE, PS and PET in routinely available FFPE human colon samples. This method combines high spatial resolution and sensitivity with non-destructive analysis, offering a comprehensive approach to study MP accumulation in human tissues. By establishing an optimized workflow preserving the colon tissue architecture, the OPTIR method offers an exclusive opportunity to identify and characterize MP particles in FFPE samples while obtaining the histopathological information required to interpret the role of these particles in the clinical context.

### Providing a clear definition of microplastic polymers after de-paraffinization

The need for deparaffinization arises because paraffin can greatly reduce the signal of substances underneath, significantly overwhelming the spectral characteristics of the embedded tissue ^44^. Sample deparaffinization might however result in residues of paraffin or polyisobutylene ^49,50^ that can challenge distinct MP particle analysis on the tissue sections. **Fig. 3** exemplifies how minor residues of paraffin and polyisobutylene remain on the sample slide adjacent to the tissue sections. Furthermore, the discrimination between PE and paraffin is challenging, given that their infrared spectra show only minor differences. However, the band at wavenumber 2959cm^-1^, indicated by a dashed line in **Fig. 3C**, allowed for a clear discrimination between the different materials.

**Fig. 3.**
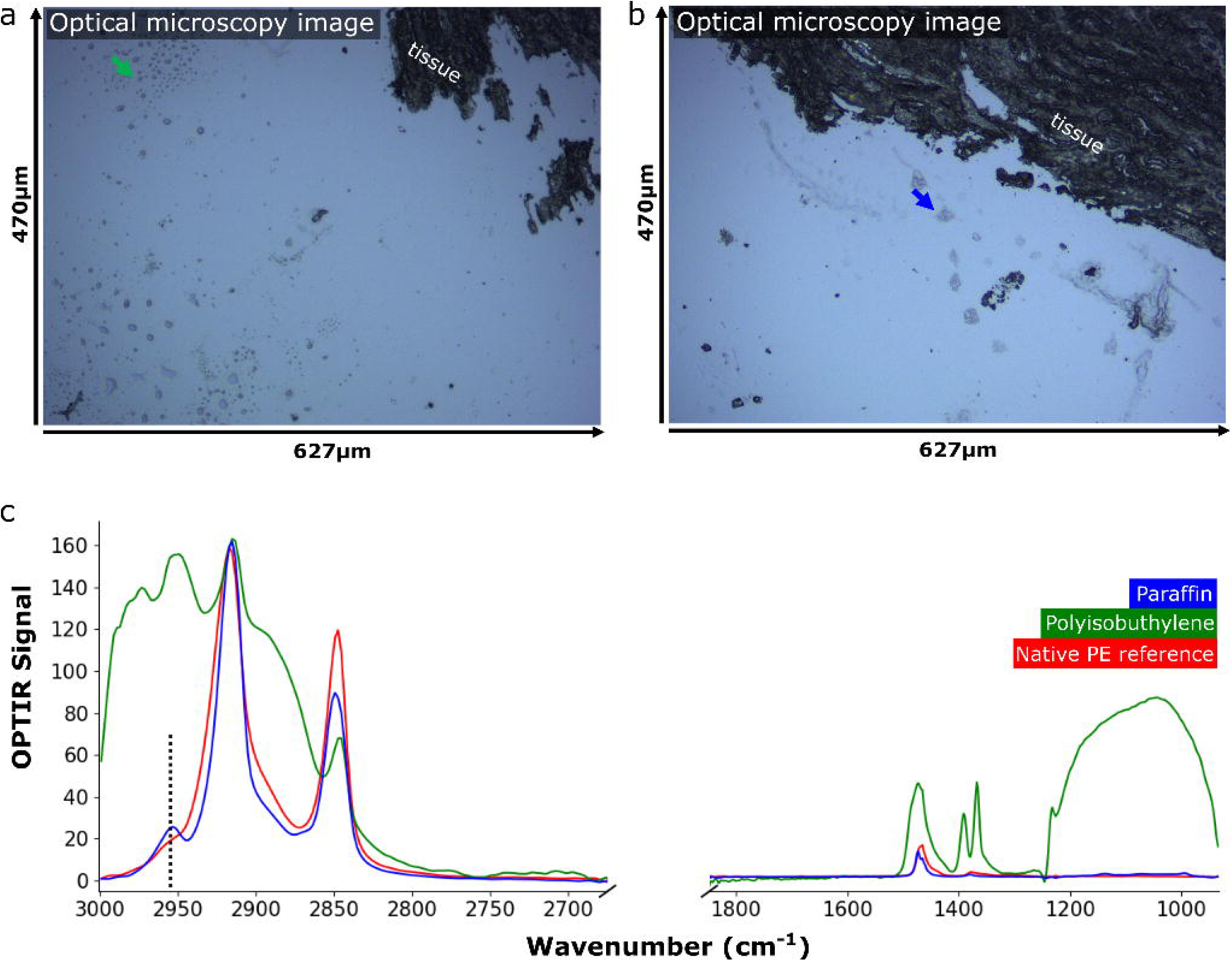
Example of material residues on a tissue slide after deparaffinization and discrimination from MP particles by exploiting characteristic spectral features. Residues of **A.** polyisobutylene (green arrow) and **B.** paraffin (blue arrow) that challenge the identification of MP particles. **C.** One specific band, centered at wavenumber 2959 cm^-1^ can be used to clearly discriminate paraffin and PE, a common MP polymer. In contrast, polyisobutylene exhibits a spectral signature that can be clearly differentiated from that of paraffin, indicating that the presence of polyisobutylene does not interfere with the detection of PE.

### Identification and localization of polyethylene (PE), polystyrene (PS) and polyethylene terephthalate (PET) polymers in distinct regions of interest

Three human colon sections (sample 1, sample 2 and sample 3) were autologously analysed in this study using the optimized workflow described above. With this, we were able to unambiguously identify PE, PS as well as PET particles. PE particles (approximately 3-6 µm in size) were detected in region I and II of sample 1 and sample 2; a detailed analysis of sample 1 resulted in twenty-one PE particles with predominantly spherical shape, thereof thirteen in region I and eight in region II. **Fig. 4** exemplarily illustrates the detection and identification of PE particles found in region II of sample 1. As demonstrated, we were able to clearly localize MP particles in organic tissue based on single-wavelength scans. For validation, full spectral analysis of the detected particles revealed the chemical fingerprint of PE and delineated the amide I and amide II bands, indicative for tissue. Notably, both amide bands exhibited a spectral shift compared to the adjacent tissue (amide I from 1660 cm^-1^ to 1645 cm^-1^ and amide II from 1545 cm^-1^ to 1563 cm^-1^), suggesting potential alterations of PE. To further validate the distinct plastic type as PE, we conducted a detailed comparison of the signal intensities in the C-H fingerprint region: Discrimination of PE from polyamide was achieved by demonstrating a significantly higher intensity of PE (2853 cm^-1^ and 2923 cm^-1^) compared to polyamide (2861 cm^-1^ and 2929 cm^-1^) ^51^. In infrared spectroscopy, spectral shifts might result from changes in the vibrational frequencies due to the interaction of the polymer with the surrounding matrix. In the present example, such a shift in the amide bands can be further associated with aging effects of MP particles ^52^. According to a study by Fernández-Gonzáles et al., a typical aging effect for PE is the emergence of an OH-band at around 1640 cm^-1^; since this band is located precisely in the region of the amide I band, the overlap of these two bands may result in a seemingly shifted amide I band in the final spectrum ^52^. However, these processes can be complex and are currently under investigation. **Fig. 5** highlights the observation of an isolated PS particle (approximately 10 µm in size), detected in region I of sample 2. The differences between the ratio plots for PS and PE (**Fig. 5**) are not as strong as compared to those used for validating PE particles (**Fig. 4**). This is because PS has non-zero absorption at 2855 cm^-1^ (visible in the plotted PS spectrum), which contributes to the appearance of signals in the PE ratio, as the wavenumber 2855 cm^-1^ is used for this ratio plot. However, PS can distinctively be observed via direct comparison of the calculated ratio plots. This example nicely illustrates a weakness of pure ratio plots, thus full spectral analysis of the sample of interest is recommended to obtain comprehensive chemical information. Detailed analysis of the individual PS MP particle further revealed distinct amide bands without any shift (amide I band at 1660 cm^-1^, amide II band at 1540 cm^-1^). However, a significant difference was observed in the appearance of a prominent band at the wavenumber 1735 cm^-1^, which is not present in the reference spectrum of native PS. However, it is important to note that this study does not investigate any comparison with native PE, PS or PET particles that have been subjected to the same chemical treatments as the tissue sections. Consequently, this band may originate from a change in the chemical composition of the particle. Another explanation for the appearance of the band at 1735 cm^-1^ might be the presence of lipids attached to the MP particle. **Fig. 6** illustrates a PET based fiber (approximately 1000µm x 13µm) which was identified within region I of sample 3. The ratio plot indicative for PET reveals the distinct position of the detected fiber and illustrates its embedding within the tissue matrix. Upon closer examination by a high-resolution chemical image in **Fig. 6B**, variations in the signal intensities can be observed at multiple locations alongside the fiber. Full spectral analysis of these specific areas revealed the incorporation of the fiber into the surrounding tissue matrix^53^.

**Fig. 4.**
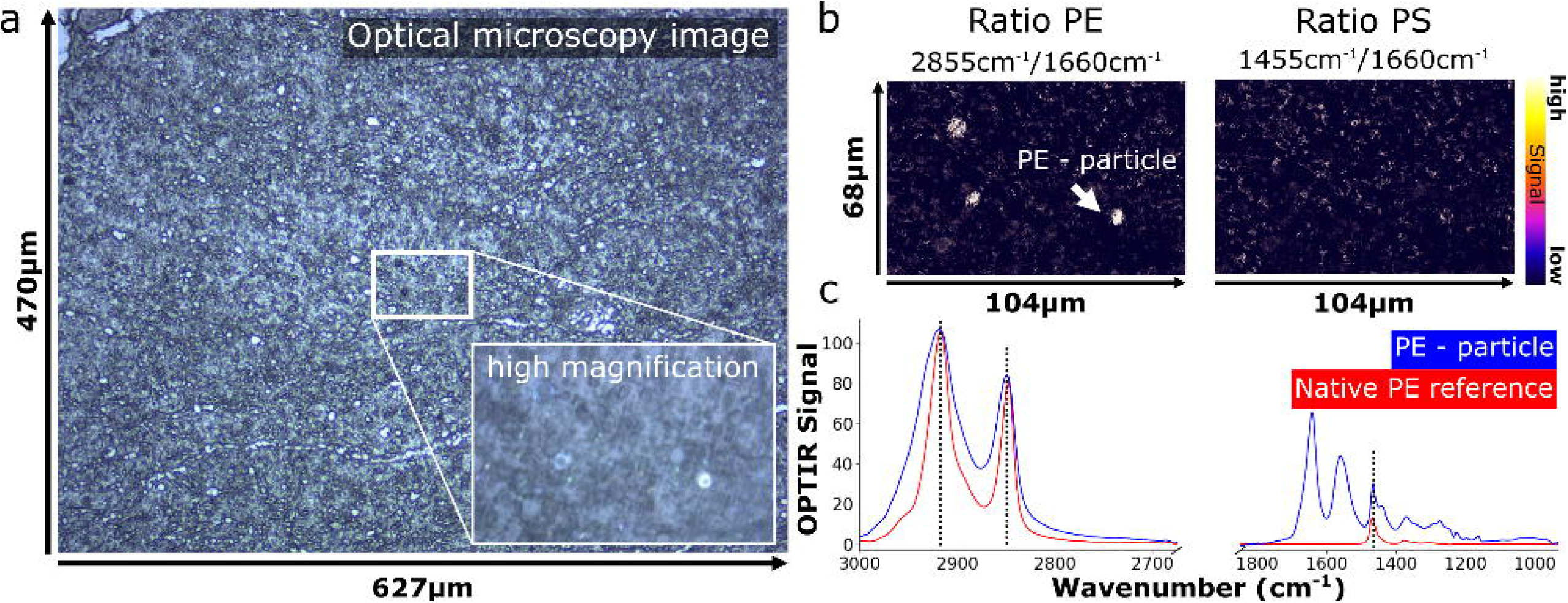
Validation of identified polyethylene particles based on their spectral features. **A.** Visible image of region II (muscularis) in sample 1 of the FFPE colon tissue sections; here, three out of the eight identified particles are displayed as bright spots in a chemical ratio image (104 x 68 µm). **B.** Ratio image for PS serving as negative control, indicating that the detected MP particles are PE. **C.** The infrared plot shows a comparison between the reference spectrum of PE (red) and the spectrum of the identified PE particle (blue) enabling validation of the identified PE particle (correlation of characteristic PE bands, observed at wavenumbers 2923 cm^-1^, 2855 cm^-1^ and 1467 cm^-1^).

**Fig. 5.**
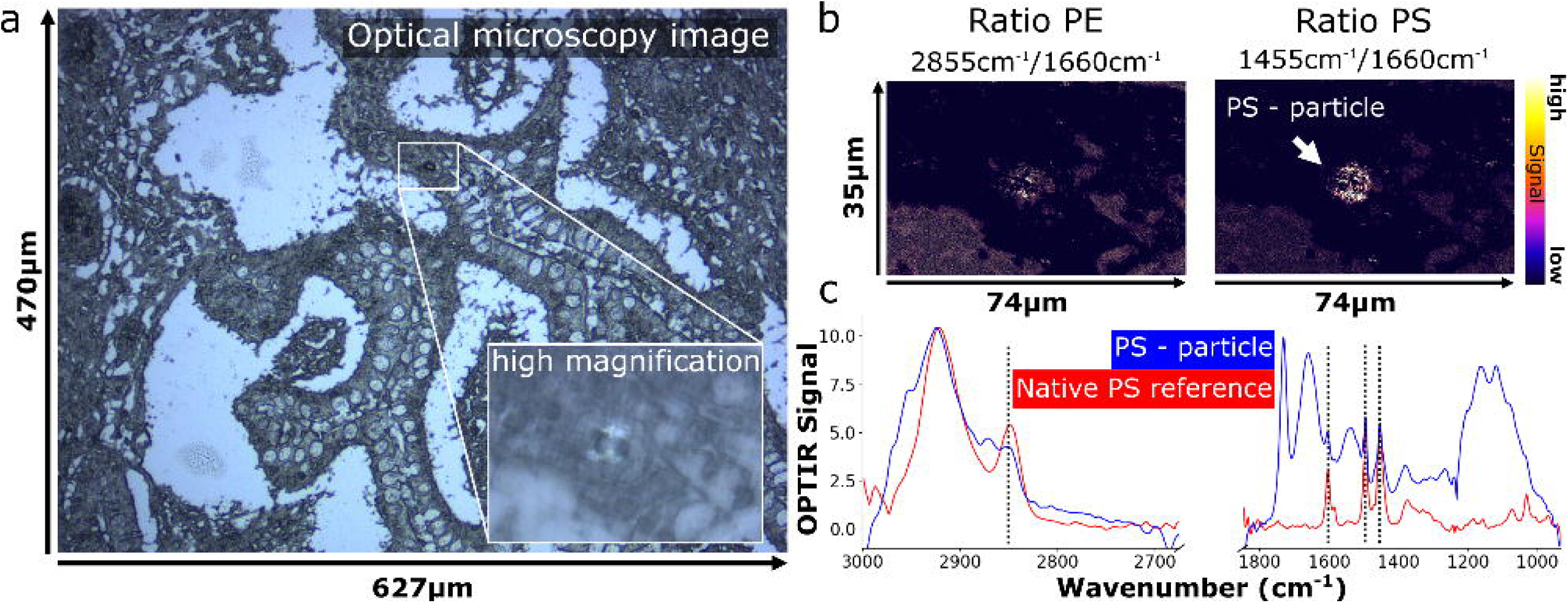
Validation of identified polystyrene particles based on their spectral features. A. Visible image of region I (mucosa) in sample 2 of the FFPE colon tissue sections; here, one particle was identified and displayed as bright spot in a chemical ratio image (74 x 35 µm). **B.** Ratio image for PE serving as negative control, indicating that the detected MP particles are PS. **C.** The infrared plot shows a comparison between the reference spectrum of PS (red) and the spectrum of the identified PS particle (blue) enabling validation of the identified PS particle (correlation of characteristic PS bands, observed at wavenumbers 1602 cm^-1^, 1495 cm^-1^ and 1455 cm^-1^).

**Fig. 6.**
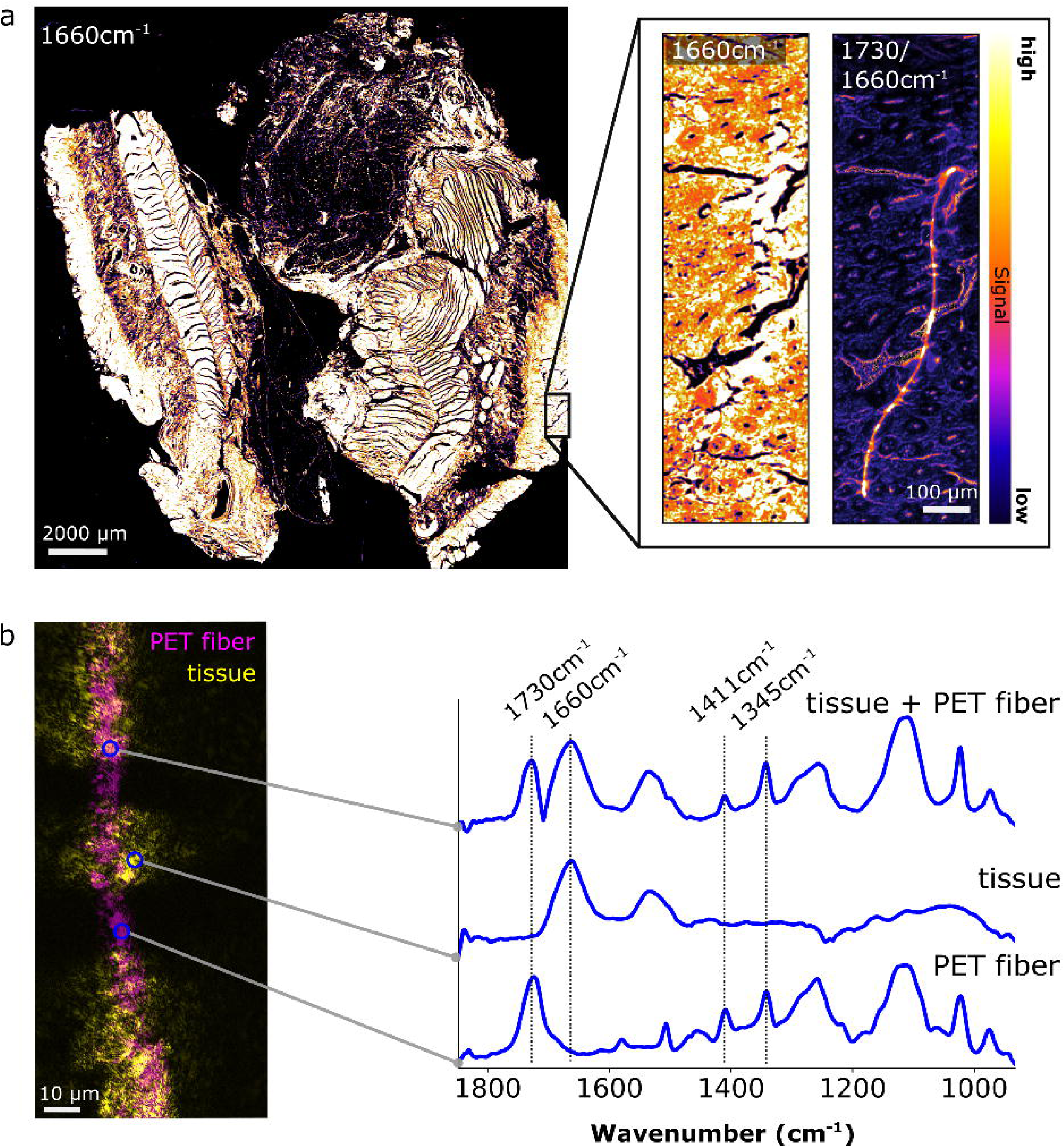
Validation of identified polyethylene terephthalate (PET) fiber based on its spectral features. **A.** Chemical image (recorded at wavenumber 1660 cm^-1^) of sample 3 provides an overview of the entire cross-section (LDIR, 3 µm resolution). Subsequent chemical imaging of a defined area with higher spatial resolution (LDIR, 1 µm resolution) results in a clear visualization of the fiber. The calculated ratio image (wavenumber 1730/1660 cm^-1^) strongly indicates that the PET fiber is embedded within the tissue matrix. **B.** The chemical image overlay (1660cm^-1^ and 1730 cm^-1^) reveals the distinct spatial distribution of the fiber, tissue material, and areas where the fiber is fully embedded. Full spectral analysis at each point is presented in the spectral fingerprint region (OPTIR, 500 nm resolution).

### Spatial localization of particles using a hyperspectral data cube

In addition to wavelength-specific chemical images (e.g. ratio plots) and full spectra at single spots of interest, the acquisition of hyperspectral images offers a more comprehensive view of the scene. A hyperspectral image (HSI, also referred to as hyperspectral data cube) contains a full spectrum for each image pixel. Such a multidimensional data set enables application of advanced multivariate and machine learning based methods for identification and characterization of different materials or substances embedded in a complex matrix. **Fig. 7** exemplarily shows a hyperspectral image with a pixel size of 500 nm at an optical resolution of ∼500 nm of the PE particle marked in blue in **Fig. 4**. The distinct delineation of the PE particle is visualized (**Fig. 7A**) as the image was extracted from the hyperspectral cube based on the characteristic wavenumber for PE at 2855 cm^-1^. HSI data enables more precise determination of the boundaries of the particle and its location within the tissue. In contrast, the second image (**Fig. 7B**) serves as a negative control as it highlights the surrounding tissue, by extracting the OPTIR signal of the amide I band at 1660 cm^-1^. The region where the particle is located exhibits a similarly high signal intensity, as indicated by the bright areas. This observation supports the conclusion that the investigated particle is embedded within the tissue matrix.

**Fig. 7.**
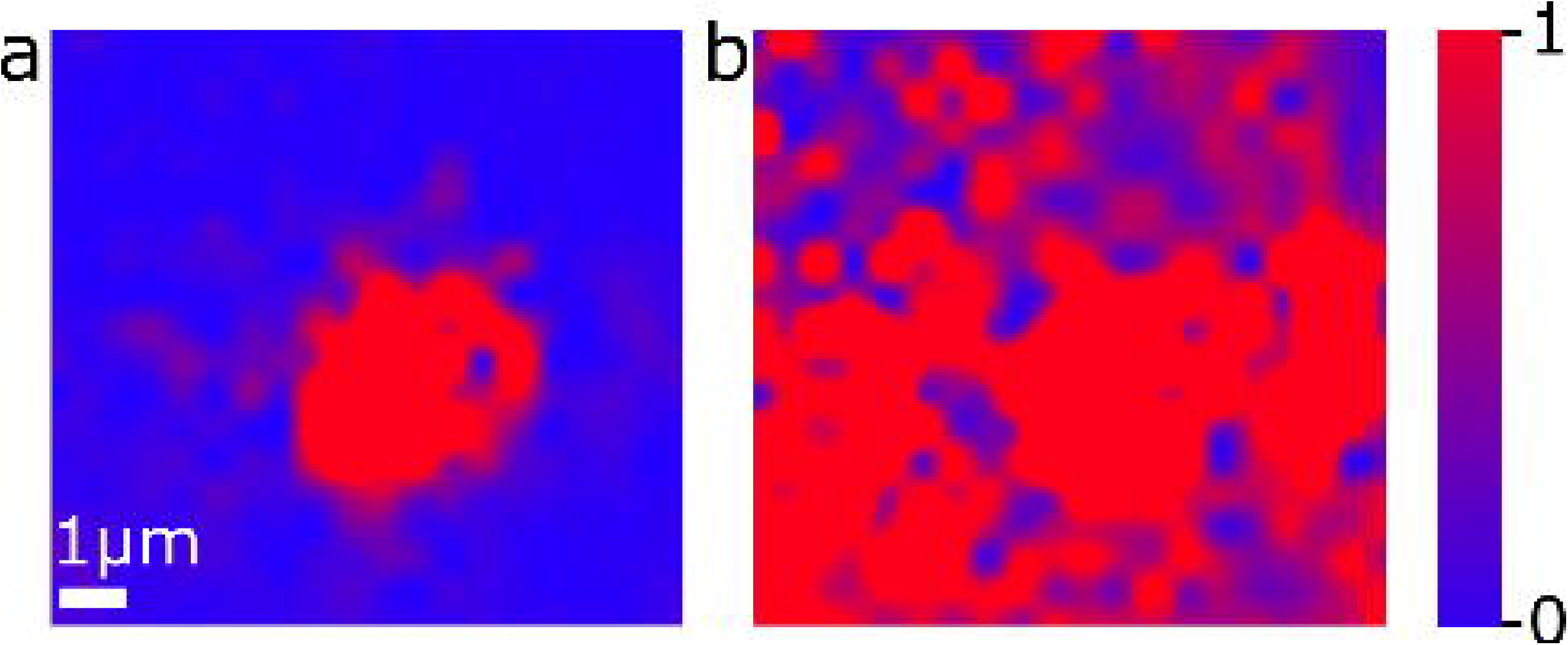
In-depth characterization and spatial localization of suspicious particles by recording a hyperspectral data cube. By exemplarily using the PE particle marked in blue in Fig. 4, a hyperspectral image (HSI) was generated (pixel step size: 500 nm, optical resolution: ∼500 nm). For the HSI, a full infrared spectrum was acquired for each displayed pixel, subsequently two images were extracted from this hyperspectral data cube: **A.** One highlighting PE (2855 cm^-1^) and **B.** one highlighting matrix material represented by the amide I band (1660 cm^-1^). The region where the particle is located exhibits a similarly high signal intensity as the surrounding area. This indicates the presence of biological tissue, leading to the conclusion that the investigated particle is embedded within the biological tissue matrix.

### Inflammatory lesions in colon tissue detected by H&E staining

The H&E stained regions showed the precise spatial particle distribution of MPs in close association with the characteristic histopathological features of inflammation (**Fig. 8**). In particular, MPs were localized near focal, patchy lesions, areas where lymphocytic infiltration selectively affects certain regions while sparing others, similar to colitis. In addition, MPs were identified in regions that showed thickening of the muscularis propria, where we also noted significant neural hyperplasia, characterized by an increased number and size of nerve bundles. These features - patchy chronic inflammation, transmural thickening, and neural hyperplasia observed in the vicinity of MPs - might suggest a possible link between MP exposure and colon inflammation.

**Fig. 8.**
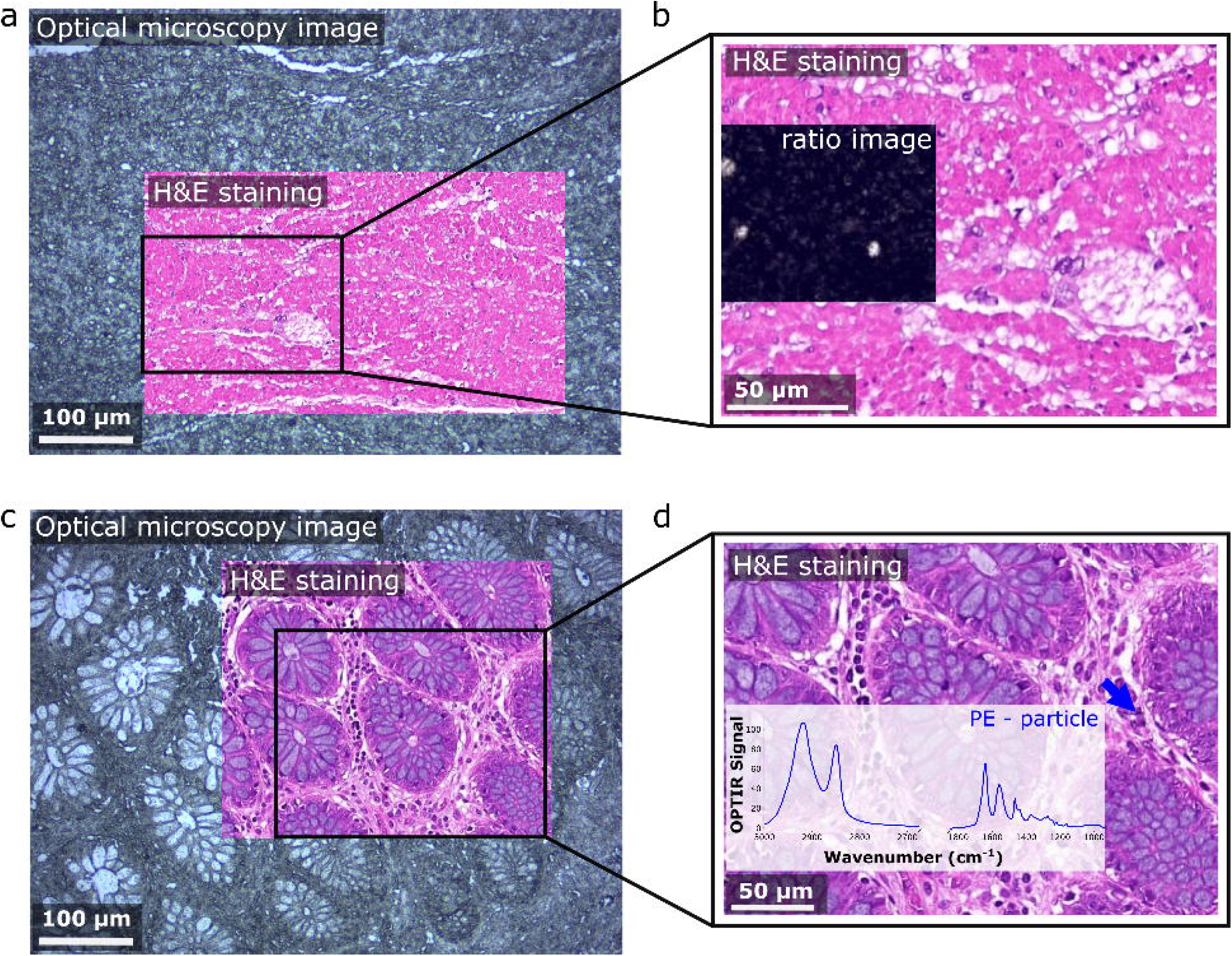
H&E analysis **A**. Optical image of sample 1 in region II (muscularis) and a partial overlay with the corresponding H&E image. **B.** Magnified view of the H&E stained region where PE particles were found and verified via chemical image analysis. **C.** Optical image of sample 1 in region I (mucosa) and a partial overlay with the corresponding H&E image. **D.** Depicts a magnified view of the H&E stained region; an isolated PE particle found in that region and verified by chemical analysis is indicated with a blue arrow. The corresponding spectrum of this particle is shown as inset.

According to the evident accumulation of MP particles in human tissues ^8–29^, the need for further research into potential health threats is of utmost importance. Although several experimental approaches have been introduced ^48^, the clear identification of MP particles remains a technical challenge, especially in terms of spatial resolution when FTIR microscopes are used. The here introduced OPTIR method represents a highly promising approach, as it surpasses the diffraction limit, thus enabling reliable measurements down to the submicron range. This non-contact technique is ideal for chemical imaging of soft tissues. It is non-destructive and allows for subsequent analysis or histopathological staining of samples. The measurement quality rivals that of FTIR-ATR, without the need for specialized substrates or sample preparation, thus preserving standardized protocols already well-integrated into clinical routines.

## Summary and Conclusion

In summary, our study pioneers the detection and characterization of MP particles in FFPE routine human colon tissue samples and is also the first to detect the presence of MPs in this clinical context. By utilizing innovative optical photothermal infrared spectroscopy (OPTIR), we achieved unprecedented spatial resolution that surpasses the limits of conventional FTIR microscopy and enables the precise identification of polyethylene (PE), polystyrene (PS) and polyethylene terephthalate (PET) particles. This non-destructive method preserves tissue structure, allowing subsequent histopathological analyses that are crucial for understanding the clinical impact of MP accumulation. For in-depth results, an automated quantification method for particles has yet to be developed. As demonstrated in the illustrated infrared spectra, this method is also affected by the chemical influence of the surrounding tissue. In contrast, methods involving tissue ^2,5,9,10,35,40,41,45,49,52^ ^11–16,33,36,44,47,48^ eliminate this effect and receive a clear spectrum of the polymers. The strong presence of amide bands I and II, indicative of biological components, results in a mixed spectrum of tissue and polymer, complicating the chemical identification of a polymer type. Furthermore, alterations in the polymer composition caused by environmental influences or chemical agents employed for tissue fixation constitute a critical factor in the identification process of the polymer type. Degradation of polymers may result in the appearance of new spectral peaks, shifting of peaks, or the disappearance of existing ones. Given that the tissue sections investigated are untreated samples, there is no available information regarding the origin or age of these particles. This may explain the minor spectral variations observed when compared to the reference spectra.

Our results show a significant correlation between the presence of MP and characteristic histopathological features of colitis. These histomorphological alterations suggest that MPs may play a role in possibly inducing an inflammatory process.

To conclude, this pioneering study presents a new approach to the detection of MPs in clinical FFPE tissue samples, providing both chemical and histopathological insights. The correlation between MPs and characteristic signs of colitis suggests a possible link between plastic exposure and disease exacerbation. This work lays the groundwork for future investigations into the broader health effects of MPs, particularly in the context of inflammatory diseases such as colitis.

## ETHICS DECLARATION

Conflict of interest: The authors have not disclosed any competing interests.

## Data Availability

Enquiries about data availability should be directed to the authors.

## FUNDING

V.K., K.D., M.S.B., T.L., L.K., M.B. acknowledges the support from microONE, a COMET Modul under the lead of CBmed GmbH, which is funded by the federal ministries BMK and BMDW, the provinces of Styria and Vienna, and managed by the Austrian Research Promotion Agency (FFG) within the COMET—Competence Centers for Excellent Technologies—program. V.K., K.D., M.S.B. and M.B. further acknowledge support by research subsidies granted by the government of Upper Austria (Grant Nr. FTI 2022 (HIQUAMP): Wi-2021-303205/13-Au). Financial support was also received from the Austrian Federal Ministry of Science, Research and Economy, the National Foundation for Research, Technology and Development, and the Christian Doppler Research Association, as well as Siemens Healthineers for their financial and scientific support. L.K. was also supported by a European Union Horizon 2020 Marie Sklodowska-Curie Doctoral Network grants (ALKATRAS, n. 675712; FANTOM, n. P101072735 and eRaDicate, n. 101119427) as well as BM Fonds (n. 15142), the Margaretha Hehberger Stiftung (n. 15142), the Christian-Doppler Lab for Applied Metabolomics (CDL-AM), and the Austrian Science Fund (grants FWF: P26011, P29251, P 34781 as well as the International PhD Program in Translational Oncology IPPTO 59.doc.funds). Additionally, this research was funded by the Vienna Science and Technology Fund (WWTF), grant number LS19-018, and L.K. received support from a European Union Horizon 2020 Marie Sklodowska-Curie Innovative Training Network grant (ALKATRAS). L.K. is a member of the European Research Initiative for ALK-Related Malignancies (www.erialcl.net).

## CONFLICT OF INTEREST

The authors have not disclosed any competing interests.

## AUTHOR CONTRIBUTIONS

Conceptualization: E.S.G., T.L., L.K.; methodology: E.S.G., V.K., D.K., T.L., L.K., M.B.; software: not applicable; validation: E.S.G., V.K., D.K., M.S.B., B.T., L.K., M.B.; formal analysis: E.S.G., V.K., M.S., L.K., M.B.; investigation: E.S.G., V.K., D.K., M.S.B., M.S., L.K., M.B.; resources: L.K.; data curation: E.S.G..; writing—original draft: E.S.G., V.K., M.S.; writing—review & editing: E.S.G., V.K., D.K., T.L., B.T., L.K., M.B.; visualization: V.K.; supervision: E.S.G., B.T., L.K., M.B.; project administration: E.S.G., M.B.; funding: L.K. All authors have read and agreed to the published version of the manuscript.

## Notes

### Competing Interest Statement

The authors have declared no competing interest.

### Author Declarations

Ethical approval for the study was given by the local ethics committee (Medical University Vienna, Vienna, Austria, No. 1003/2024). Informed consent was waived off.

